# Direct detection of SARS-CoV-2 RNA using high-contrast pH-sensitive dyes

**DOI:** 10.1101/2020.12.26.20248878

**Authors:** Timothy A. Brown, Katherine S. Schaefer, Arthur Tsang, Hyun Ah Yi, Jonathan B. Grimm, Andrew L. Lemire, Fadi M. Jradi, Charles Kim, Kevin McGowan, Kimberly Ritola, Derek T. Armstrong, Heba H. Mostafa, Wyatt Korff, Ronald D. Vale, Luke D. Lavis

## Abstract

The worldwide COVID-19 pandemic has had devastating effects on health, healthcare infrastructure, social structure, and economics. One of the limiting factors in containing the spread of this virus has been the lack of widespread availability of fast, inexpensive, and reliable methods for testing of individuals. Frequent screening for infected and often asymptomatic people is a cornerstone of pandemic management plans. Here, we introduce two pH sensitive ‘LAMPshade’ dyes as novel readouts in an isothermal RT-LAMP amplification assay for SARS-CoV-2 RNA. The resulting JaneliaLAMP (jLAMP) assay is robust, simple, inexpensive, has low technical requirements and we describe its use and performance in direct testing of contrived and clinical samples without RNA extraction.

## Introduction

In the wake of the COVID-19 pandemic, there has been a significant worldwide effort toward the development of tests that effectively identify SARS-CoV-2-infected individuals. Management strategies for the COVID-19 outbreak are often centered on the ability to conduct widespread testing for the SARS-CoV-2 virus. Quantitative RT-PCR (qRT-PCR) is the gold standard assay for virus detection, as it is quantitative, robust, and uses equipment and protocols common in clinical testing laboratories. qRT-PCR assays are best suited for use in centralized testing facilities designed to handle large numbers of patient samples shipped from different sampling locations. This process requires transport logistics, expensive equipment, and technical expertise, however, which can lead to long turnaround times for test results. In addition, reliance on a single assay places pressure on the supply chain for test components, allowing possible disruptions under high demands. Finally, there is a need for more rapid and inexpensive tests that could be applied at point-of-care (POC), near-patient venues, or in surveillance testing. To diversify the testing arsenal and to enable alternative testing modalities, we have developed JaneliaLAMP or “jLAMP”; a RT-LAMP assay for the SARS-CoV2 viral RNA which is inexpensive and simple to perform.

The LAMP technique is an isothermal nucleic acid amplification method developed 20 years ago^1^, and has been used to detect a variety of human pathogens^2^. LAMP typically uses a set of 6 primers which create a dumbbell-shaped single DNA strand with end loops created by self-annealing segments. This substrate is then amplified by other primers using a strand-displacing thermostable DNA polymerase in an isothermal reaction. The method has been adapted to include reverse transcriptase (RT) to create the cDNA LAMP template from RNA viruses (**Figure 1a**)^3^. The LAMP reaction produces large amounts of DNA and a molecule of pyrophosphate and a proton are released for each nucleobase added in the DNA synthesis process (**Figure 1a**). All three of these LAMP products have been used to determine the progress of the reaction.^1,4^ Protons can be detected by using weakly buffered conditions, which allows a large decrease in pH to accompany DNA synthesis; this can be read out by colorimetric dyes as pioneered by Tanner *et al*^5^. Here, we introduce two novel high-contrast pH indicators, called ‘LAMPshade’ dyes, and demonstrate their use in jLAMP assays for the detection of SARS-CoV-2 viral RNA. These dyes can be used as both colorimetric and fluorescent read-outs. Using inactivated SARS-COV-2 virus and clinical samples, we show that the jLAMP assay does not require RNA extraction, performing well in a direct testing protocol from common viral transport media. We demonstrate sensitivity, specificity, stability, and validation using clinical nasopharyngeal swab samples and extend the jLAMP protocol to direct testing from saliva.

**Figure 1.**
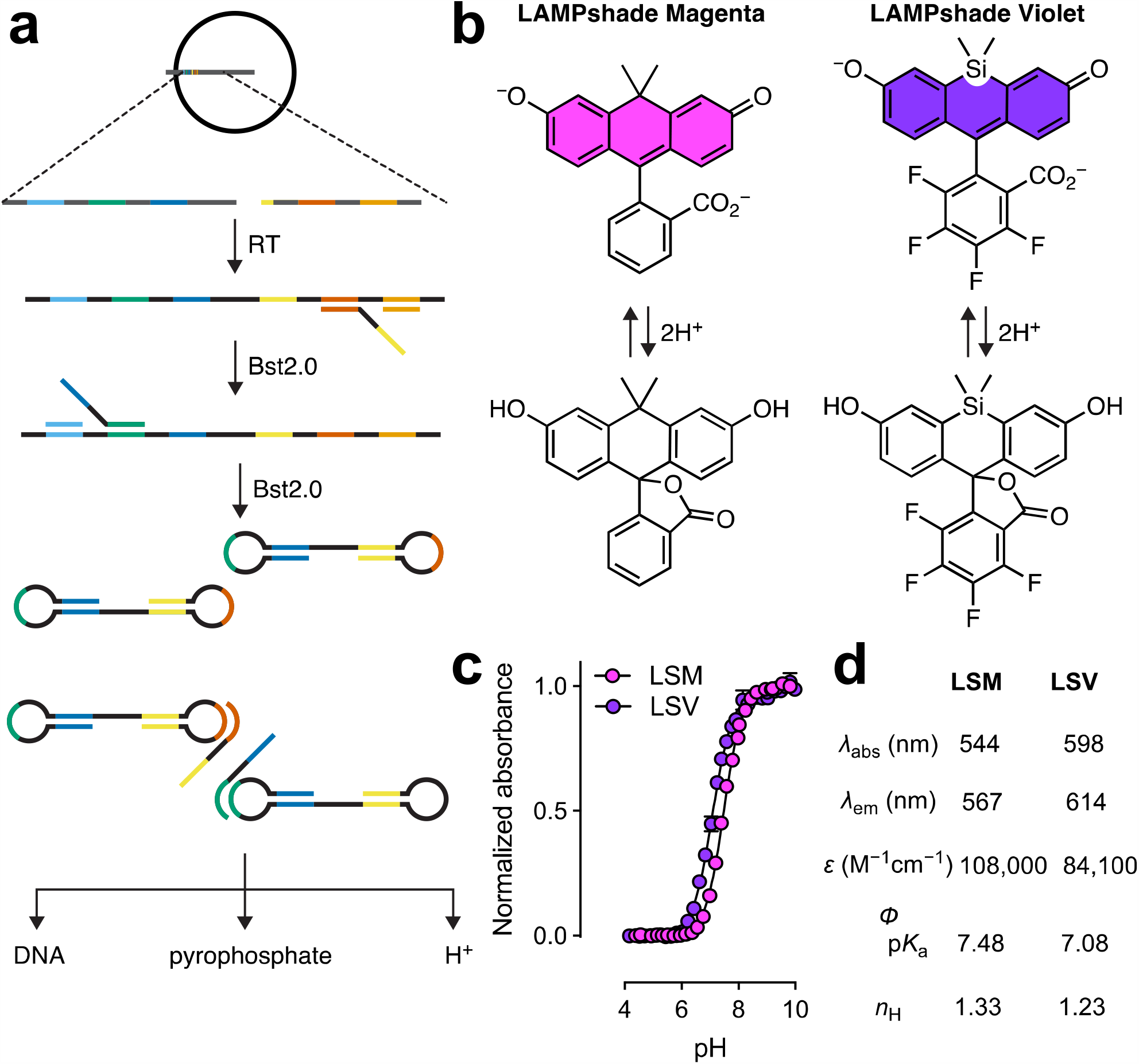
LAMP and LAMPshade dyes. (**a**) Schematic of LAMP assays for detection of SARS-CoV-2. Bst 2.0 is a strand-displacing DNA polymerase (NEB) (**b**) structures of SiFl598 and CFl. (**c**) pH fluorescence response curves. (**d**) Dye photo properties including maximum excitation and emission wavelengths, extinction coefficient, quantum yield, pKa, and Hill coefficient.

## Results

### Use of Novel pH-indicator LAMPshade dyes

We screened a panel of possible pH sensitive dyes based on carbofluorescein (CFl) and Si-fluorescein (SiFl) for their utility in LAMP assays. We identified two compounds, which showed optimal contrast and perfomance: carbofluorescein (LAMPshade Magenta) and 4,5,6,7-tetrafluoro-Si-fluorescein (LAMPshade Violet **Figure 1b-d**)^6,7^. Examination of the pH titration of these dyes reveal they both exhibit optimal p*K*_a_ values (p*K*_a_ = 7.08-7.48) for LAMP reactions performed under low buffering capacity, which are initiated at ∼pH 8.5 and are complete at ∼pH 6.0 ^5^. In addition, both LAMPshade dyes exhibit a cooperative transition between a highly colored dianionic species and a colorless lactone form, resulting in a steep colorimetric/fluorometric pH dependent transition, reflected in their Hill coefficients. We compared the pH sensitivity of the LAMPshade dyes to the common pH indicator Phenol Red, which has been used for visual detection of LAMP reactions and is a component in commercial assay kits (**Figure 2a**). Aqueous solutions of the LAMPshade dyes are highly colored above their p*K*_a_ but are colorless below with a relatively steep transition. In contrast, Phenol Red changes from red to yellow as pH decreases. The Phenol Red ratiometric readout and shallower transition can lead to ambiguous determinations, especially using visual readout with colorblind individuals. We then compared these dyes in LAMP assays (**Figure 2b**) using weakly buffered conditions. In line with the pH titration, the LAMPshade dyes offer a greater contrast due to their binary response and steeper color transition. In addition to yielding colorimetric readouts by eye, the LAMPshade dyes can be visualized using fluorescence excitation with a hand-held UV light (**Figure 2c**) or measured in a fluorescence plate reader, a quantitative PCR instrument, or other photo-detecting instrument. Here, we chose to focus on colorimetric readout to minimize technical barriers.

**Figure 2.**
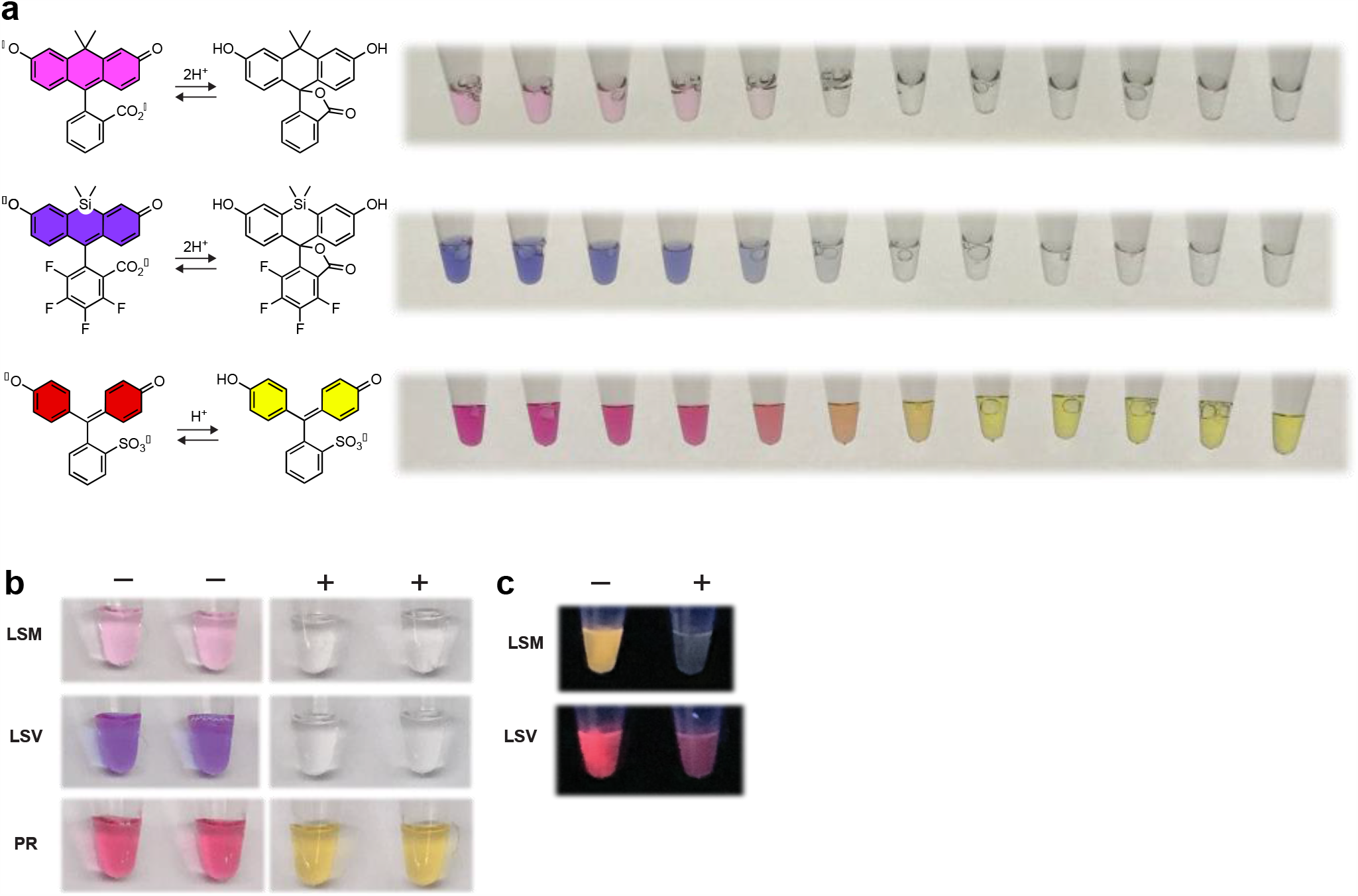
Colorimetric dye pH response and LAMP readouts with various pH indicators. (**a**) Images of solutions of LAMPshade Magenta, LAMPshade Violet, and Phenol Red at varying pH values ranging from pH 9.0 to 6.25 in 0.25 Unit increments, left to right. (**b**) Images of negative (-) and positive (+) jLAMP reactions under normal light. (**c**) Images under hand-held UV (365 nm) light using LAMPshade Magenta and Violet.

### LAMP Primers and Reaction Optimization

Having established the utility of the LAMPshade dyes in the LAMP assay, we focused on optimizing the reaction for SARS-CoV-2 detection. We envisioned jLAMP assay workflow as displayed in **Figure 3a**, where heat treated swab media or saliva is directly added to the jLAMP master mix, heated at 60 °C and read out visually. First, we screened several sets of LAMP primers^8–10^ to achieve optimum performance and found that those designed by D. Wang^10^ yielded the best sensitivity. We also designed primers specific for the human RPP30 and 18S rRNA genes to be used to test sample integrity. Second, we considered the buffer composition as commercial assay kits use proprietary formulations. Buffer choice is influenced by many factors, including the transport media used for the sample and the specific p*K*_a_ of the LAMPshade dye. After numerous buffer iterations, we found 1-2 mM Tris-HCl buffer as an acceptable starting concentration; higher concentrations (>3mM) resulted in reactions that were slower to change the indicator color. The pH tuning of these buffers for various viral transport media is shown in **Figure 3b**. Finally, we optimized the assay duration as LAMP primers can form non-template-mediated self-constructions given enough time, starting the exponential amplification. We defined the endpoint of the assay as the time at least 30 minutes prior to the typical conversion of non-template control sample; endpoints ranged 30-60 min, depending on the primer set.

**Figure 3.**
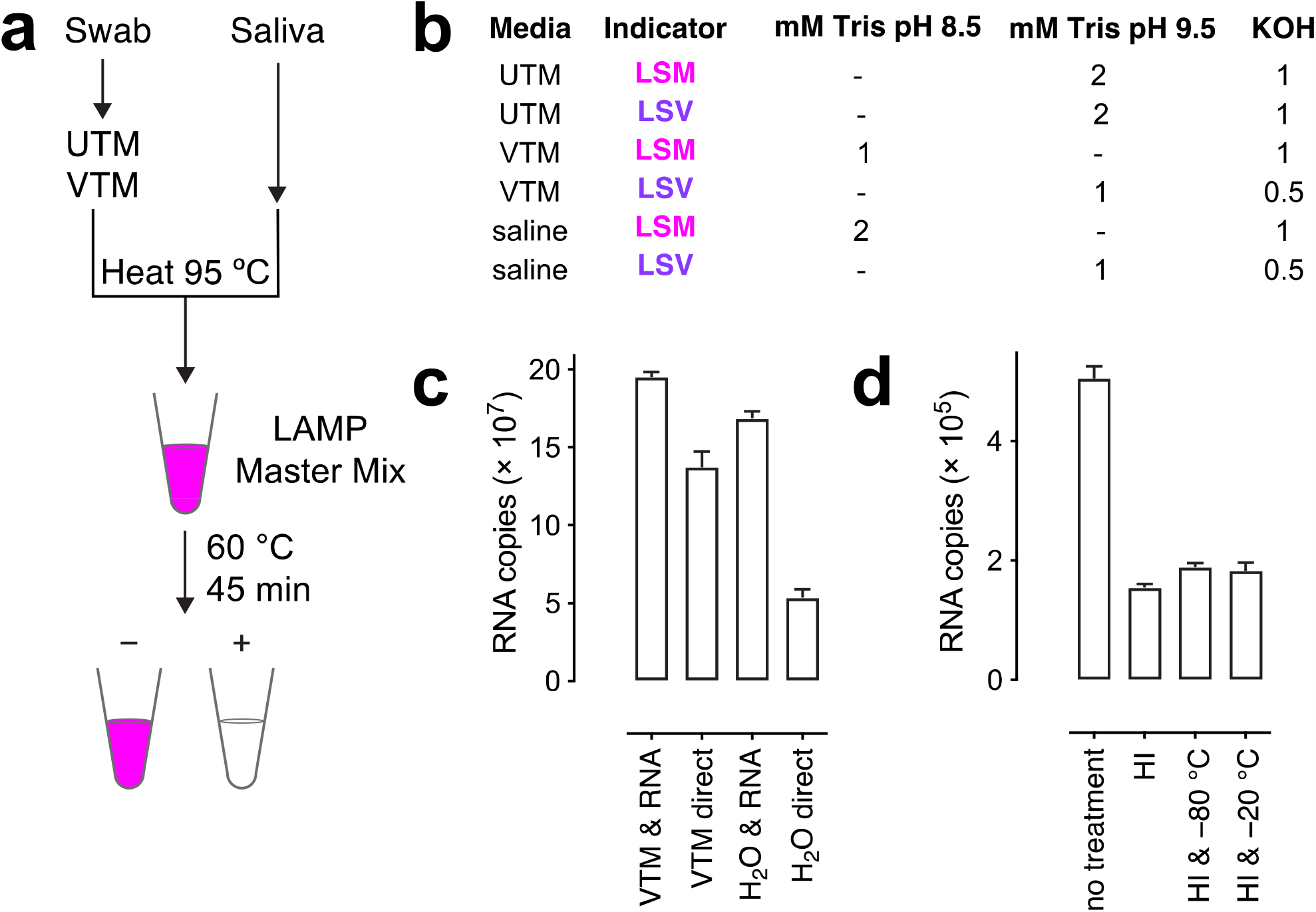
jLAMP assay optimization. (**a**) Workflow of jLAMP assay. (**b**) Variable buffer composition optimizations for viral transport media. (**c**) Comparison of HCoV229E viral RNA detected after RNA purification or directly from samples in either VTM or H_2_O. (**d**) Effects of heat and freeze-thaw treatments on RNA recovery in direct qRT-PCR sample assays.

### Effect of virus sample handling and processing in direct qRT-PCR

RNA purification from patient samples is commonly employed in qRT-PCR and other nucleic acid assays. To reduce cost and complexity, we sought to define conditions where the jLAMP assay could function without RNA extraction, using direct addition of sample to the reaction. As a proxy for SARS-CoV-2, we used the non-lethal human coronavirus 229E (HCoV 229E) strain to test for the availability of viral RNA using qRT-PCR under different conditions. The propagated HCoV 229E was diluted and spiked into viral transport media (VTM) or deionized water. Samples were either processed by RNA extraction followed by qRT-PCR or directly used in qRT-PCR. Direct assay of HCoV 229E in VTM yielded 70% of the RNA extracted sample (**Figure 3c**). RNA detection from HCoV 229E in water was only 31% of the RNA extracted control. Additional experiments indicated that serum proteins in VTM are responsible for the increased recoveries relative to water (data not shown). Assuming that HCoV229E is similar to SARS-CoV-2 we inferred from these data that direct testing of viral samples without RNA extraction was feasible with a modest, acceptable loss of sensitivity.

In addition to RNA extraction, clinical COVID-19 samples are often heat inactivated to limit viral exposure during handling and then stored frozen. We tested the effects of a heat inactivation and a single freeze/thaw cycle. HCoV229E was spiked into VTM and divided into four tubes. Each tube was further exposed to heat inactivation at 65 °C for 30 min and different storage conditions as shown **Figure 3d**. Heat inactivation decreased the available RNA to about 33% of the control but additional freeze/thaw treatment did not further decrease the recovery. Thus, employing both freeze/thaw and viral heat inactivation in the testing workflow is reasonable, but does further affect sensitivity.

### Limits of Detection and False Positives in viral sample media

Although our RT-LAMP assay was initially developed with synthetic RNA templates, we found that the most useful biomimetic was to test various media that had been spiked with known amounts of heat-inactivated SARS-CoV-2 virus (HI-SARS-CoV-2). **Figure 4a** displays data compiled from multiple independent experiments to determine assay sensitivity in various CDC/WHO approved nasal swab media used in sample collection from COVID-19 patients. This includes universal transport media (UTM), viral transport media (VTM) and normal saline (0.9% w/w). Clean sample media was spiked with various dilutions of HI-SARS-CoV-2 virus (ATCC) and tested directly, without RNA purification or processing. The 95% limit of detection (LoD) is defined in the FDA’s emergency use authorization (EUA) guidelines as that RNA copy number that is detected at 95% frequency or better from at least 20 samples. UTM is somewhat inhibitory relative to VTM and saline with 95% LoD values of 90.6, 45.3, and 22.6 RNA copies, respectively. The lowest LoDs are 11.3, 2.8, and 2.8 respectively. UTM shows a more defined detection cutoff below 11.3 RNA copies per reaction as shown in **Figure 4b**.

**Figure 4.**
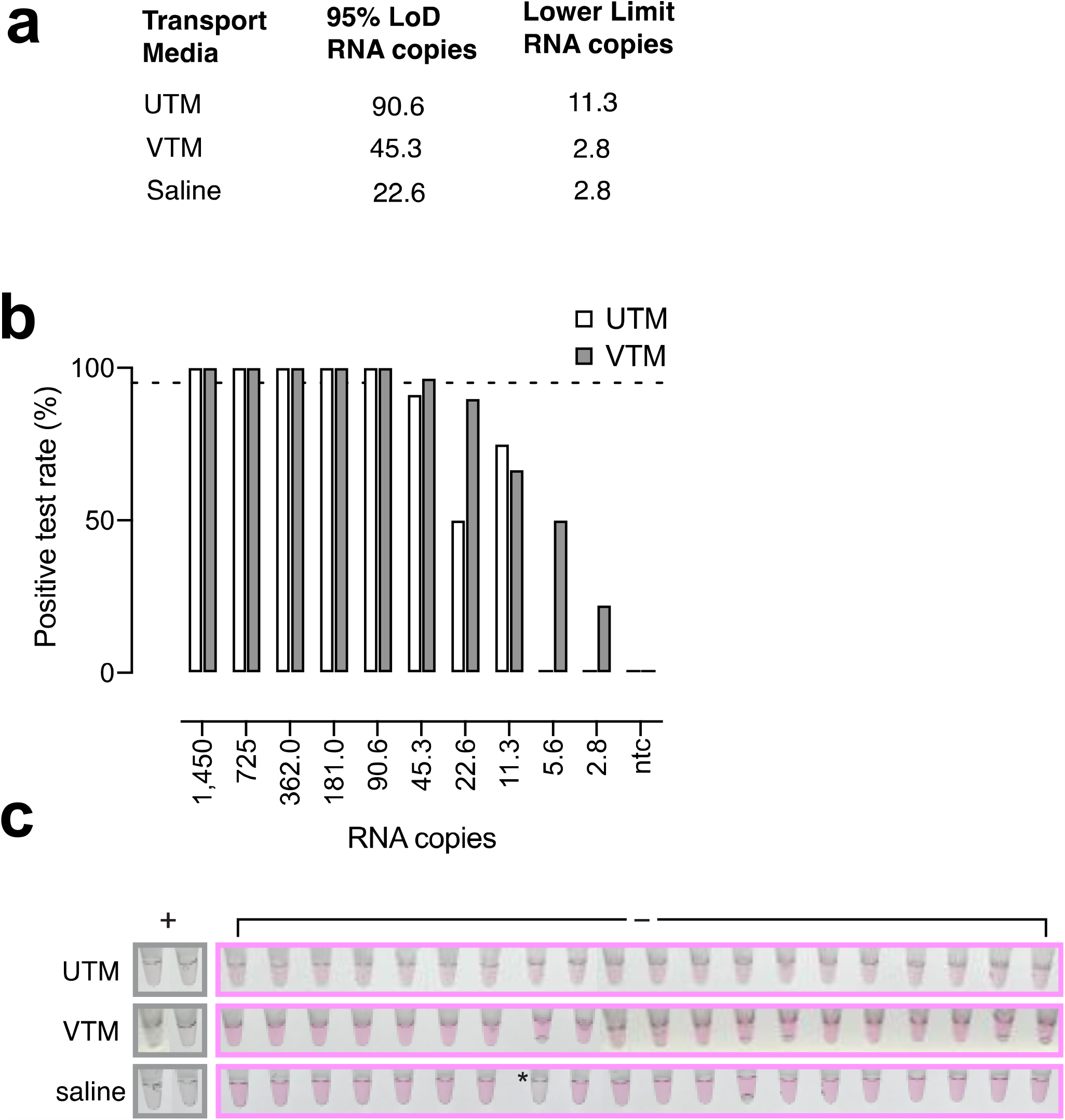
Sensitivity and specificity of jLAMP. (**a**) Limits of detection of jLAMP in various viral transport media spiked with HI-SARS-CoV-2. Data was compiled from multiple independent experiments. (**b**) jLAMP sensitivity in UTM and VTM at low viral copy numbers. **c**) False-positive rate in different swab sample transport media: VTM, UTM, or saline. Positive controls (+) contained heat inactivated SARS-CoV-2 virus at ∼260 copies. The single false positive in saline is marked with an asterisk.

LAMP reactions can yield false positive results since there is a small but significant probability for some combination of the six primers to self-assemble into non-specific primer-only DNA amplifications. The frequency of these events increases with incubation time. As this is an endpoint assay, we have chosen that stop time to balance high sensitivity with low false positives. To validate our choice of a 45 min incubation time using the DW primers, we tested the frequency of non-template constructions using clean virus sample buffers. Twenty replicates for each of the three samples buffers were run without template (**Figure 4c**). Only one tube of the 60 replicates turned clear, representing a false positive rate of 1.66%. As with all LAMP-based assays, increasing the assay time beyond this endpoint is expected to increase sensitivity at the cost of greater false positivity.

### LAMP sensitivity in spiked negative nasopharyngeal swab samples

We obtained clinical nasopharyngeal (NP) swab samples in UTM which were identified as negative for SARS-CoV-2 RNA by RT-PCR. A subset of these samples (*n* = 13) were spiked with serial dilutions of HI-SARS-CoV-2 virus and assayed to determine the degree of inhibition that might be contributed by native RNases, mucus, and other substances from nasal secretions transferred into the swab media. **Figure 5a** shows the percent detection at each of the SARS-CoV-2 dilution values. Although in this specific experiment we did not achieve the 95% LoD of 90.6 RNA copies, there was minimal inhibition relative to clean UTM (**Figure 4b**) and we could detect lower amounts of RNA with modest reproducibility; detecting 5.6 RNA copies and 2.8 RNA copies in 38.5% and 30.8% of the samples, respectively.

**Figure 5.**
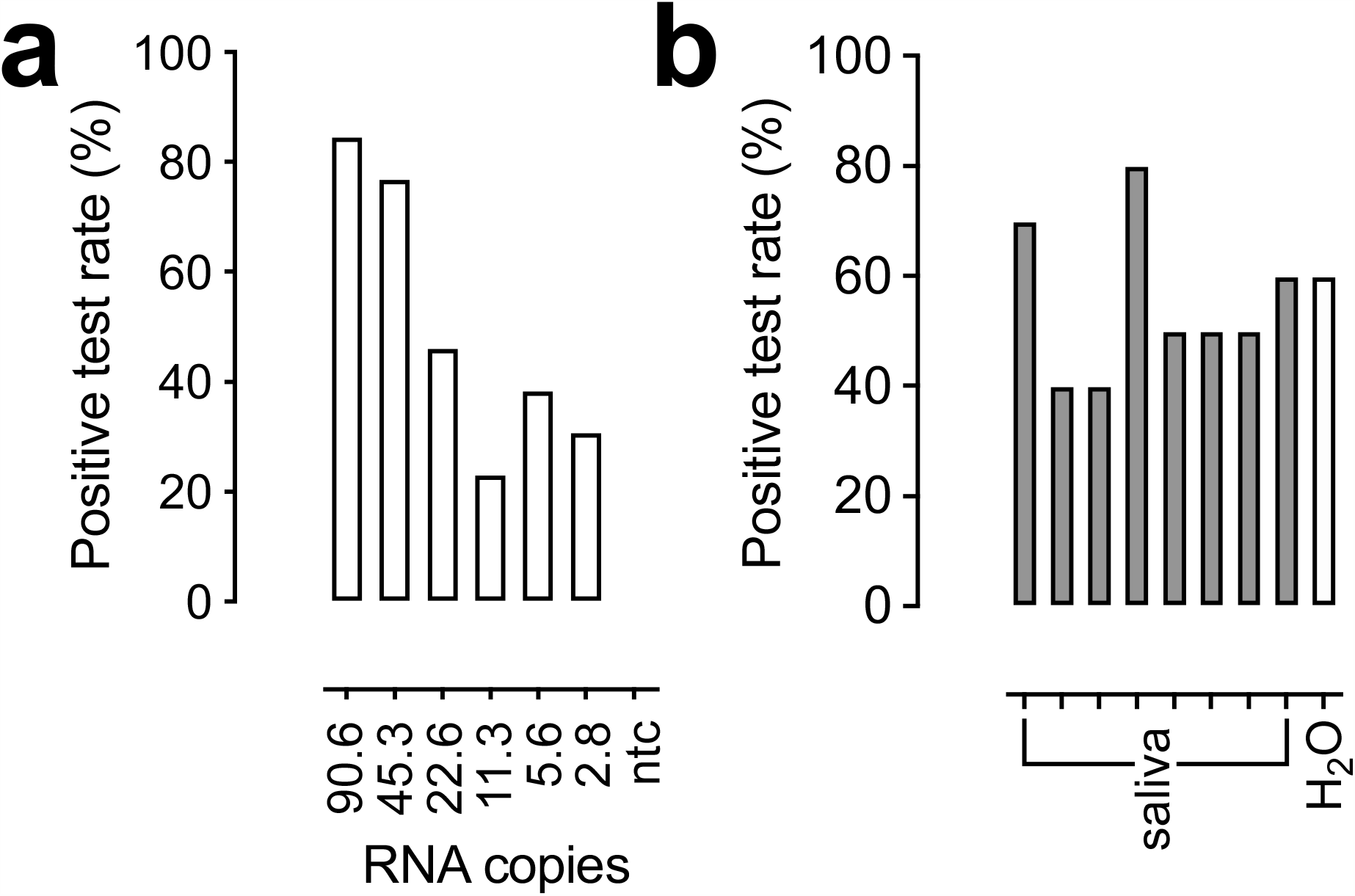
jLAMP assay optimization. (**a**) jLAMP sensitivity in COVID-19 negative NP swab samples. Thirteen COVID-19 negative samples were spiked with serially diluted heat inactivated SARS-CoV-2. Percent positive samples are plotted against the corresponding RNA copy values; LAMP reactions were performed with LAMPshade Magenta. (**b**) Saliva sample testing for LAMP performance. Eight normal saliva samples were spiked with 23 copies of heat inactivated SARS-CoV-2 (n = 10). Percent positive for each saliva sample is displayed relative to a water control.

### Preliminary validation of LAMP using direct saliva samples and sputum

To assess the feasibility of performing the LAMP assay directly on saliva samples, we ran several experiments using normal saliva spiked with HI-SARS-CoV-2. Untreated saliva was inhibitory and showed a large variation between samples. During initial trials using proteases to degrade RNAses, we found that treating samples at 95 °C for 10 min needed for protease inactivation achieved the same results with or without protease. **Figure 5b** shows that after heat treating saliva from 8 individuals and a subsequent challenge with 23 viral copies, all samples supported LAMP detection, albeit at varying detection efficiencies. Sputum samples were also tested after treatment with DTT and diluted prior to spiking with HI-SARS-CoV-2 virus. The LAMP assay failed to detect viral RNA in these samples (data not shown).

### LAMP performance with direct assay of COVID-19 patient NP swab samples

We then compared the efficacy of the LAMP assay with a qRT-PCR using clinical samples. We used COVID-19 samples from patients that were designated as positive using a sensitive two-gene qRT-PCR assay (Altona)^11^. In this initial diagnostic PCR assay, a portion of the swab sample was subjected to RNA extraction protocol, concentrated 10-fold after RNA extraction, and a 10 µL RNA volume was added to the qRT-PCR master mix. In contrast, the jLAMP assay uses 2 µL of direct sample, circumventing the RNA extraction and concentration steps. We expected lower sensitivity relative to the qRT-PCR as the jLAMP assay uses 50× less starting material than the qRT-PCR assay, with further loss of sensitivity due to the use of UTM (**Figure 4b**) and other factors mentioned above. **Figure 6** displays jLAMP test results from each of the COVID-19 positive patient samples against the corresponding Ct values from a second qRT-PCR assay using the CDC N-gene primer set. The jLAMP successfully detected RNA in 23 of the 30 original positive samples. Sensitivity tapered off below 157 RNA copies, while still detecting about 17.4 copies (Ct=36.2); the assay failed to detect genome numbers below 10 copies in these patient samples. We note that four of LAMP-negative samples were also negative using CDC N-gene qRT-PCR, likely due to the lack of RNA concentration after purification, the lowered amount of test volume, or the heat inactivation of these samples. We also tested 30 negative COVID-19 patient samples; all but one tested negative in the LAMP reactions. Interestingly, this solitary sample was retested using qRT-PCR and found to be a false negative (Ct=37.2; data not shown).

**Figure 6.**
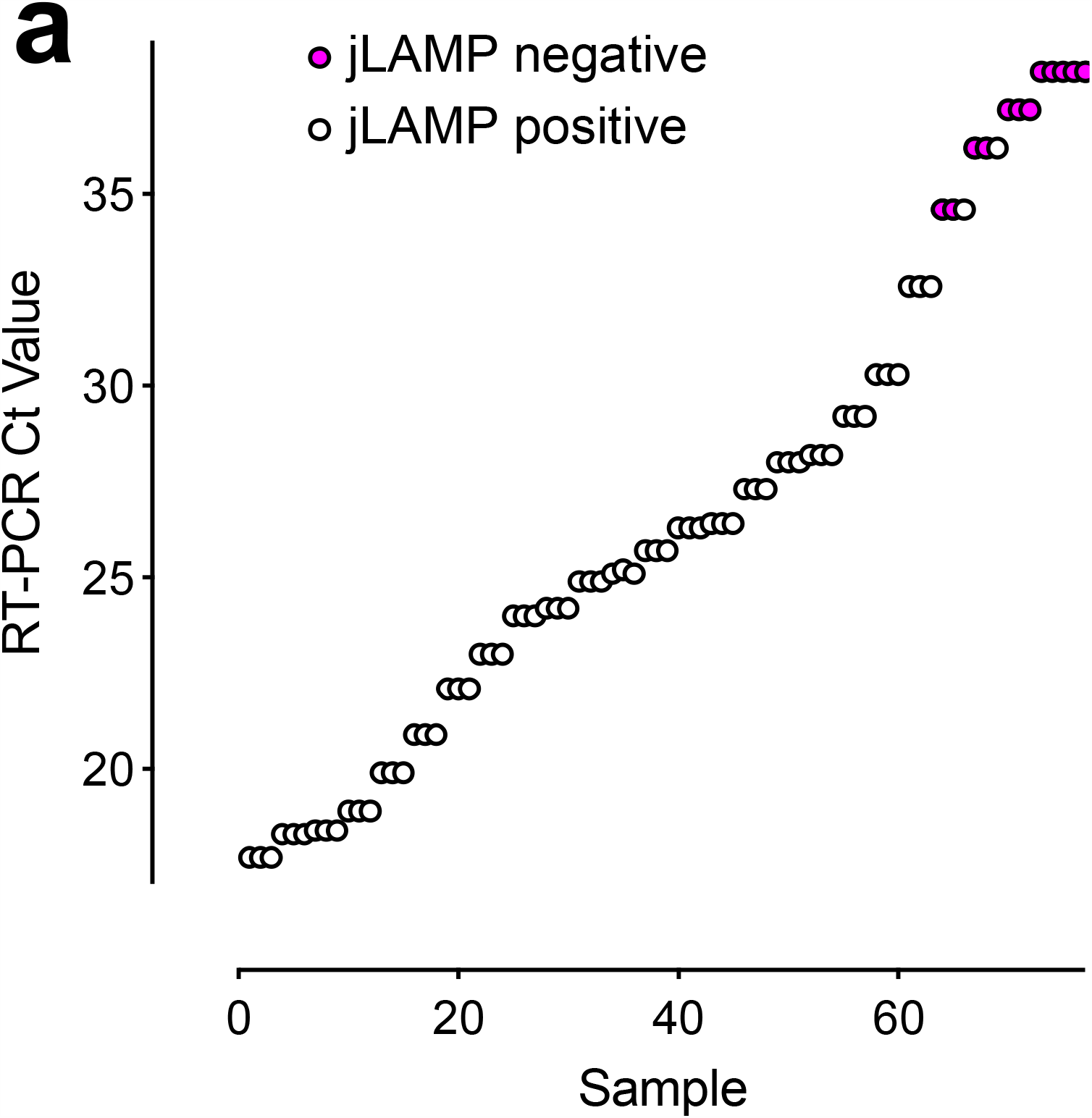
**(a)** COVID-19 patient nasal swab quantification and jLAMP test results. SARS-CoV-2 positive nasopharyngeal swabs in UTM were LAMP tested in triplicate (*n* = 30). qRT-PCR Ct values obtained from the CDC N-gene primer set are reported based on assays after heat inactivation. LAMP negative samples are displayed in red text.

### LAMP primer specificity

We conducted an *in silico* analysis to determine the organism exclusivity and specificity of the DW LAMP primer set. We aligned the primer sequences to 41,858 high-quality genomes from the NCBI Virus SARS-CoV-2 data hub that contained the LAMP assay primer region in the N gene from RefSeq NC_045512.2 (see Methods). The sequence identity for all six LAMP primers in this alignment was 99.8% or greater, and the per-base mismatches for the last five nucleotides of each primer are quantified in Figure 7a. The greatest frequency of mismatches is within the FIP primer at the −4 position, which should be well tolerated. It is therefore likely that this primer set will detect all sequence variants of SARS-CoV-2 (as of December 9, 2020) although some variants may amplify with less efficiency. However, all 6 LAMP primers also have partial or complete sequence identity with the SARS coronavirus ZJ01. This implies that patients with SARS-CoV from the original pandemic in 2003 could test positive with this assay. Only one primer (Wang LB) had partial sequence identity with MERS-CoV, which is insufficient to generate LAMP products. We confirmed the *in silico* analysis using the LAMP primer set using RNA from SARS-CoV and MERS-CoV (**Figure 7b**). Finally, to control for false positives stemming from other organisms expected to be present within patient swabs, we used a respiratory control panel of bacteria and viruses obtained from Zeptometrix (**Figure 7c**). Control #1 includes adenovirus type 1, adenovirus type 3, adenovirus type 31, *C. pneumoniae*, influenza A H1N1 pdm, influenza H3N2, metapneumovirus type 8, *M. pneumoniae*, parainfluenza type 1, parainfluenza type 4, and rhinovirus type 1A. Control #2 includes *B. parapertussis, B. pertussis*, coronavirus HKU-1, coronavirus NL63, coronavirus OC43, influenza A H1N1, influenza B, parainfluenza type 2, parainfluenza type 3 and RSV type A. The LAMP assay was negative with these control panels, yielding no cross-specificity from these pathogens.

**Figure 7.**
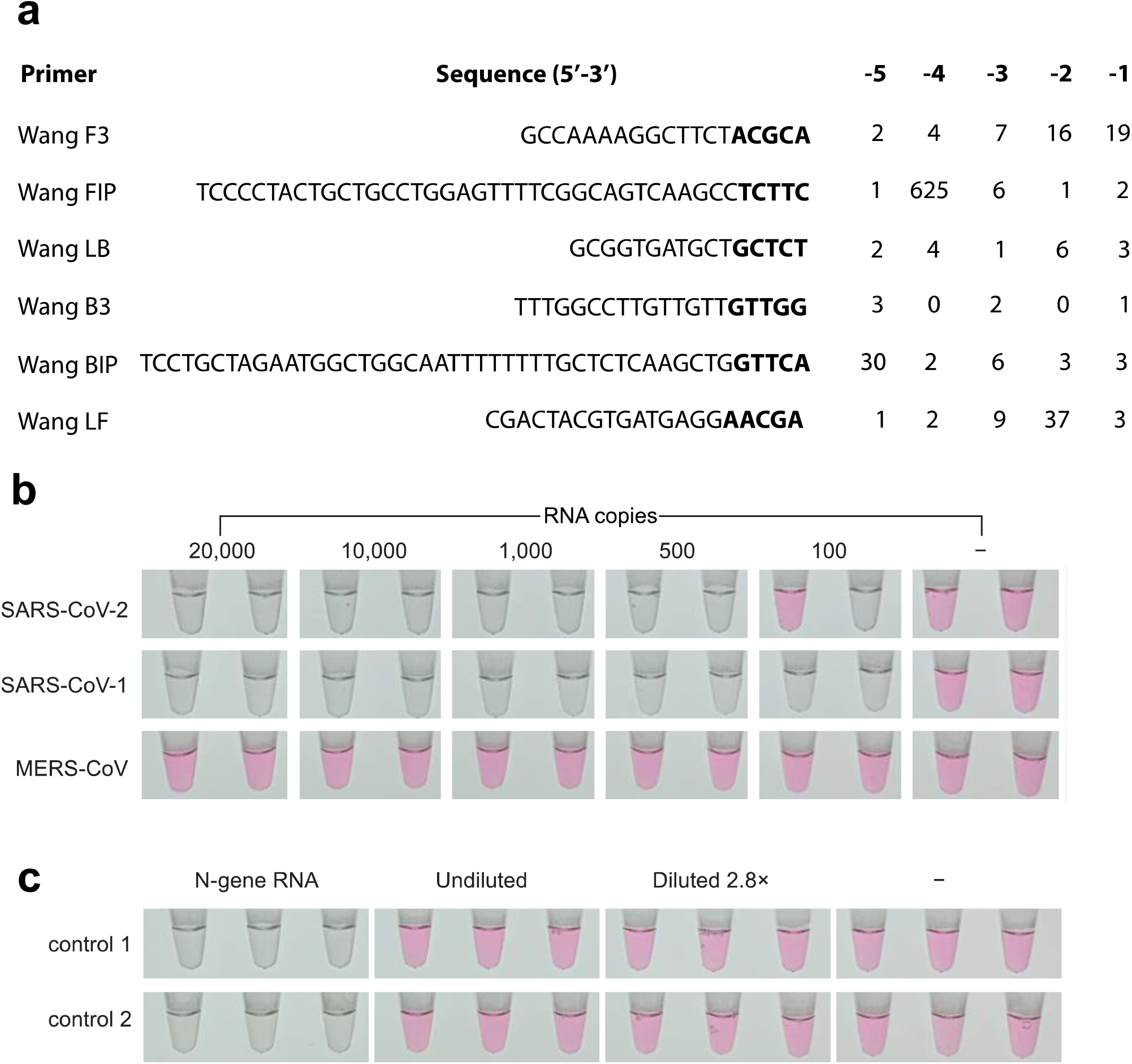
jLAMP specificity and exclusivity. (**a**) Number of per base mismatches to published SARS-CoV-2 genomes. 41,858 high quality SARS-CoV-2 genomves from NCBI were used to quantify per-base mismatches for the last 5 bases in each LAMP primerThe −5 position is the first bold text base and the −1 position is the last. 617 of the 625 mismatches at FIP −4 are C->A mutants (S183Y). (**b**) jLAMP results after using varying copies of SARS-CoV-2, SARS-CoV, and MERS-CoV shown in duplicate. (**c**) jLAMP reactions with RNA purified from Zeptometrix respiratory control #1 (iv) and #2 (v) to determine exclusivity. The N-gene positive control, undiluted Zeptometrix panel RNA, diluted panel RNA, and no template controls (NTC) are shown in triplicate.

### LAMP master mix stability

To simplify deployment logistics, we sought to create a stable master mix of our LAMP components to which only test sample is applied. The master mix could then be prepared and frozen for later use at a near-patient testing site. We screened a panel of 11 cryoprotectants, testing both assay interference and cryoprotective ability. The panel included various disaccharides (sucrose, maltose, trehalose), glycerol, PEG400. PEG200, proline, ethylene glycol, lithium sulfate, sodium malonate, and 1-methyl 2,4 pentanediol at various concentrations. Surprisingly, we found the least assay interference and best stability in formulations that had no cryoprotectant. The master mix is stable for at least 4 weeks when stored at −20 °C without loss of sensitivity (**Figure 8a**).

**Figure 8.**
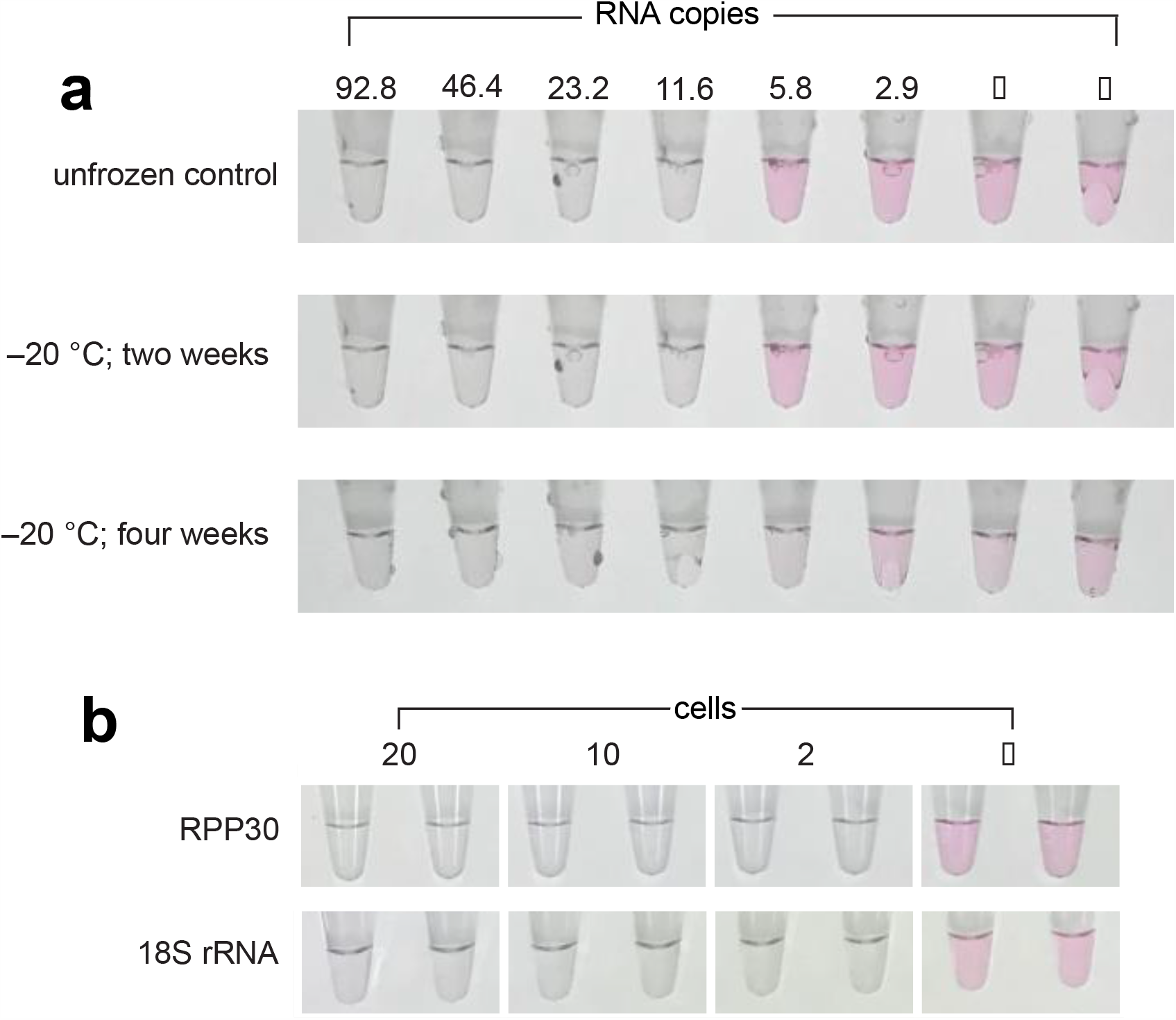
LAMP master mix storage stability and integrity controls. (**a**) Tube labels with two-fold serial dilutions of SARS-CoV-2 from 92.8 to 2.9 copies: top: unfrozen control; middle: −20 °C storage for 2 weeks; bottom: −20 °C storage for 4 weeks. (**b**) LAMP assays for RPP30 and 18S rRNA as sample integrity controls. RPP30 (RNaseP subunit) and 18S rRNA LAMP primers were tested on HEK293 mammalian cells at 20, 10, and 2 cells per tube in duplicate.

### LAMP with sample integrity control primers for RNAseP (RPP30) and 18S rRNA

To determine RNA presence and integrity in COVID-19 patient samples, an appropriate control assay should be applied to all negative sample reads. The human RPP30 and 18S rRNA genes are highly expressed and have been used as sequence targets for sample integrity tests. We tested sets of LAMP primers for both RPP30 and 18S rRNA and validated them using human HEK293 cells as a biomimic for patient cells transferred from nasal swabs. **Figure 8b** shows that both of these primer sets can detect as low as two HEK293 cells per assay. We quantified the RNA transcript level in HEK293 cells using qRT-PCR and determined that 2 cells contain an approximately 19 copies of RPP30 RNA.

## Discussion

Robust, accurate, and inexpensive COVID-19 diagnostic tests are a cornerstone of pandemic management strategies. RT-LAMP is a simple viable alternative to the standard qRT-PCR and can fill the need for a complementary COVID-19 test in near-patient venues without the need for expensive instruments and highly trained personnel. In addition, RT-LAMP can be less expensive, and does not compete with the qRT-PCR reagent supply chain. We describe here the jLAMP assay which uses novel colorimetric pH indicators, LAMPshade Violet and LAMPshade Magenta. The jLAMP assay using these dyes can be used without RNA extraction, is sensitive, specific, robust, simple, and inexpensive.

The use of pH indicators in LAMP assays was pioneered by Tanner and colleagues at New England Biolabs, who leveraged the large change in pH that occurs during the LAMP reaction from pH 8.5 to pH 6-6.5^5^. Although Phenol Red is the most popular colorimetric LAMP indicator, it has several disadvantages. First, Phenol Red has relatively high p*K*_a_ = 8.0, which is close to the starting pH of the LAMP assay. This undergoes a gradual color transition from pink above pH 8.2 to yellow below pH 6.8. It is a ratiometric dye across this range, as one color (pink) decreases as the other (yellow) increases. This can lead to ambiguous color determination, and therefore interpretation of test results. When using direct sample testing, those that are acidic can change the color of phenol red even in the absence of DNA amplification, as the *p*K_a_ and thus the color transition zone is close to the starting pH of the assay. Finally, the Phenol Red exhibits poor fluorescence properties, limiting use to colorimetric assays. In contrast, both LAMPshade dyes can be used as colorimetric or fluorescence indicators, depending on the availability of instrumentation or desire to eliminate qualitative visual readouts. Other colorimetric dyes have also been used as LAMP readouts, including leuco Crystal Violet^12^, Calcein^13^, and Hydroxynaphthol Blue^14^, which detect double-stranded DNA or magnesium. However, these dyes yield results that are arguably more ambiguous than Phenol Red or the new LAMPshade dyes.

A critical step in simplifying COVID-19 testing is avoiding the purification of RNA as the first step. This would eliminate time, cost and supply-chain bottlenecks for RNA purification reagents and kits. We quantified the loss of detection sensitivity in direct sample assays on treated samples using HCoV-229E as a proxy for SARS-CoV-2. The direct assay showed 70% sensitivity relative to the purified RNA, likely reflecting some inaccessibility of the encapsulated viral RNA in direct sampling using qRT-PCR. RNA purification also serves to inactivate the virus, allowing for increased laboratory safety. We tested a 30-minute, 65 °C heat-inactivation and achieved 33% sensitivity relative to non-heated samples. Although this heat treatment is sufficient to inactivate SARS-CoV-2^15^, it may allow degradation of RNA and loss of sensitivity. Heat treatment at 98 °C has shown to improve sensitivity in LAMP assays^16^. An additional caveat here is that HCoV-229E may not behave as SARS-CoV-2 in direct assays, as others have reported 97% sensitivity with a similar heat treatment^14^. Regardless, our data is consistent with others that direct assays and heat inactivation both support successful RT-LAMP assays, with less sensitivity relative to qRT-PCR using purified viral RNA. We consider this loss of sensitivity a reasonable trade-off in circumventing the disadvantages of RNA purification as a first step. Direct SARS-CoV-2 LAMP assays have been previously tested with varying degrees of success^16–20^.

Regarding sensitivity, we have demonstrated a 95% LoD of 22.6 copies per assay (11.3 copies/µL) and a lower limit of about 3 copies per assay (1.5 copies/µL) in normal saline (**Fig 4a**). This analytical sensitivity is essentially the same as the CDC’s 2019-nCoV Real-Time RT-PCR Diagnostic Panel with an LoD of 10 copies/µL ^21^(https://www.fda.gov/media/134922/download). Vogels and colleagues^22^ surveyed the performance of multiple qRT-PCR SARS-CoV-2 primer sets used in testing worldwide and determined that the best of these detected 1 copy/µL at 25% frequency and 10 copies/µL at 25-50% frequency. Although their assay conditions were less than optimal, the jLAMP assay described here performed similarly at these very low copy numbers (**Figure 5a**).

It should also be noted that the LoD values are analytical sensitivity measurements, and not a measure of clinical performance. In clinical samples, the jLAMP assay had a working sensitivity of about 10 copies/µL, which again is comparable to other routinely used clinical diagnostic COVID-19 tests. This level of sensitivity should be more than needed if used in a high frequency surveillance testing regimen, where sensitivity may be less consequential. Testing for clinical diagnosis may be used to manage patient care, whereas the goal of surveillance testing is to identify and sequester infectious individuals. Larremore *et al* have argued that for detection of the viral N gene RNA, those people whose samples contain less than 10,000 copies/µL are unlikely to have transmissible levels of virus ^23^. They advocate that a strategy of more frequent testing and fast reporting times largely eliminates the advantages of those tests detecting better than 1,000 copies/µL. Therefore, jLAMP vastly exceeds this criterion for use in population surveillance with frequent testing.

The specificity and exclusivity are important factors in diagnostic assays as low performance can lead to false-positive readouts. The jLAMP assay with the current primer set is predicted to detect all sequence variants of SARS-CoV-2. Accumulation of mutations in the SARS-CoV-2 population within these primer regions could render these primers dysfunctional. Fortunately, mutational mismatches in the middle or 5’ ends of the long, stable LAMP primers often have little effect on annealing and assay performance, resulting in higher mismatch tolerance than typically obtained with shorter PCR primers. Regardless, monitoring the frequency and distribution of relevant sequence variants should be an on-going effort to ensure assay integrity. We have also screened a control panel of bacteria and viruses associated with respiratory illness and determined that the jLAMP assay excludes these common organisms (**Figure 7b**). However, the primers used here do have significant homology to the original SARS-CoV virus, and we show that RNA from that virus is detected by jLAMP (**Figure 7a**). However, the exceedingly low prevalence of SARS-CoV minimizes that significance. In addition to the possibility of off-target templates, false positives also arise in LAMP reactions from primer self-constructed amplicons, analogous to primer-dimer formation in PCR. We have determined that the jLAMP yields a false positivity rate of 1.66% at our defined endpoint (**Figure 4c**). Finally, LAMP reactions can generate as much as 10-15 µg of DNA per reaction tube, which is 50 times that typically amplified in a PCR reaction. This represents a significant source of bench and instrument contamination which can also increase false positivity, and care should be taken to avoid this issue, which includes avoiding assays that require multiple additions to the assay vessel.

Our intention was to create an inexpensive assay that was simple enough to be deployed outside of a complex centralized diagnostic laboratory. To this end, LAMP assays require only a limited number of reagents and a constant heat source. Although developed with standard laboratory pipettors and benchtop thermocyclers, we have also demonstrated equivalent assay performance using an inexpensive (<$70; Amazon), and disposable glass micro pipets (Drummond). The master mix is stable at −20 °C and in the dark for at least 4 weeks and can be aliquoted into test vessels before field use. Cost and speed are also critical considerations in filling the need for frequent testing strategies. We have calculated the cost of disposable components of jLAMP to be about $2.88 per sample. That cost would double if the integrity control (RPP30 or 18S rRNA) is to be run in parallel. The assay can be completed in less than an hour from sample addition to readout, which is a considerable time savings over most PCR tests.

Although we validated the jLAMP assay using clinical nasopharyngeal swab samples, NP swabbing poses several challenges. These include swab expense and availability, the need for trained personnel, increased exposure to healthcare personnel, and discomfort to patients. Saliva sampling can circumvent many of these problems and has been shown to perform as well or better than NP swab samples in COVID-19 testing^24^. We were unable to obtain COVID-19 patient saliva samples to validate this approach using jLAMP. However, using normal saliva spiked with SARS-CoV-2, we show that the jLAMP assay can support direct testing of saliva (**Figure 5b**). The cumulative % positivity from all 8 saliva samples yields a frequency of 55% at 23 copies per assay. This is somewhat higher than the % positivity obtained from the same copy presented to normal UTM nasal swabs at 46% (**Figure 4b**). This suggests that the assay may perform as well with saliva, if not better than NP swabs in UTM. However, this is a low sample trial, and more work is needed to validate this approach using clinical saliva samples. We have collaborated with M. Rosbash laboratory at Brandeis University, who have found that some saliva samples are unsuitable for direct LAMP assays using LAMPshade Violet, and further RNA purification is needed to prevent an increase in false positives (data coordinately published).

## Materials and Methods

### LAMPshade dyes

Novel pH indicator dyes were synthesized as previously described: 4,5,6,7-tetrafluoro-Si-fluorescein (LAMPshade Violet) and carbofluorescein (CFl, LAMPshade Magenta) ^6,7^. We routinely distribute fluorescent dyes and have limited quantities available. Please contact T. Brown or L. Lavis for dye requests. Stock solutions of the LAMPshade dyes were prepared in high-quality DMSO at 1–10 mM and stored −20 °C in DMSO. These were diluted to 2.5 mM or 0.25 mM in water prior to use as 25× working stocks for a final assay concentration of 100 µM and 10 µM, respectively. Frozen stock dyes were diluted into aqueous solution fresh on the day of the assay, or within master mixes stored at −20C. We routinely keep our dyes protected from light as a general precaution against light induced changes.

### SARS-CoV-2 RT-LAMP reaction optimization

We screened LAMP primers for SARS-CoV-2 detection from prior work^8–10^ and found the best performance using those of D. Wang, which were designed to detect the viral N-gene; DW N FIP (5’-tcccctactgctgcctggagttttcggcagtcaagcctcttc-3’), DW N BIP (5’-tcctgctagaatggctggcaattttttttgctctcaagctggttca-3’), DW N F3 (5’-gccaaaaggcttctacgca-3’), DW N B3 (5’-tttggccttgttgttgttgg-3’), DW N LF (5’-cgactacgtgatgaggaacga-3’), DW N LB (5’-gcggtgatgctgctct-3’). These and other oligonucleotides were purchased from Integrated DNA Technologies.

RT-LAMP reactions contained 8 mM MgSO_4_, 30 mM KCl, 1% Tween 20, 1.6 µM or 0.8 µM FIB/BIP primers as referenced, 0.2 µM F3/B3 primers, 0.4 µM FL/BL primers, 1.4 mM each dNTP, colorimetric/fluorescent dyes as above, 7 Units of WarmStart RTx (NEB #M0380), 8 units of 120,000 Units/ml Bst 2.0 polymerase (NEB #MO537), 40 Units of NxGen RNase Inhibitor (Lucigen #30281) when working with UTM, VTM, or patient samples. The pH was adjusted depending on the sample. Figure 3b defines the appropriate buffer conditions for each type of reaction. When using VTM or water with LAMPshade violet, reactions were run using 1 mM Tris-HCl pH 9.5, and 0.5 mM KOH. To account for HEPES buffer, reactions with UTM were run using 2 mM Tris-HCl pH 9.5 and 1 mM KOH. Test samples were typically added at 2 µL, although 4 µL can be tolerated. The final reaction volume was 25 µL. The DW N-primer reactions were run at 60 °C with positive/negative readouts after 45 min. Reactions were heated using either an Applied Biosystems 2720 or GeneAmp 9700 thermocycler. Use of a water bath with mineral oil sample overlay yielded equivalent performance. Reactions were typically monitored and photographed at 15 min intervals to document reaction progress. Preliminary assay optimization was performed with quantified N-gene plasmid DNA (IDT #10006625), and in vitro transcribed RNA. However, we sought to utilize a standard that would more closely mimic clinical tests and found more consistent performance using heat inactivated SARS-CoV-2 virus (ATCC # VR-1986HK and BEI # NR-52286). Using half of the concentration of WarmStart RT (NEB) was well tolerated. Substituting MMLV-RT was not successful, but Superscript IV RT and Maxima H minus RT could feasibly be used in lieu of WarmStart RT, although with further optimization. Substitution of Bst 3.0 (NEB # M0374S) without WarmStart RT for Bst 2.0 was unsuccessful. Addition of 40 mM guanidine hydrochloride^25^ decreased the reaction time but also shortened the interval to conversion of the no template controls.

### Virus sample processing and yields

Human coronavirus 229E (HCov229E) was obtained from ATCC (VR-740) and propagated in MRC-5 cells cultured in Eagle’s Minimum Essential Medium (EMEM) supplemented with 10% fetal bovine serum (FBS) and 1× antibiotic-antimycotic solution (Gibco) at 37 °C, 5% CO2 incubator. Upon the viral infection, cells were incubated at 35 °C, 5% CO2 incubator for 2 hours. The media was replaced with EMEM with 2% FBS and 1× antibiotic-antimycotic solution and further incubated at 35 °C for 48–72 hours until cytopathic effect was exhibited. On the day of harvest, supernatant from the infected MRC-5 cells was collected. Dilute viral supernatants were used directly in spike experiments. HCoV 229E qRT-PCR standard for the viral N-gene was prepared from viral genomic RNA extracted from the original virus stock (ATCC) using QiAmp viral RNA mini kit. The cDNA was synthesized using PrimeScript 1st strand cDNA synthesis kit (Takara) followed by PCR with the following primers; HCoV229E-N-T7-fwd (5’-<u>gaaattaatacgactcactata</u>ggatggctacagtcaaatgggctgat-3’), HCoV229E-N-full-rev (5’-ttagttgacttcatcaattatgtcagt-3’). The *in vitro* transcribed RNA was prepared using the T7 RiboMAX Express System (Promega #1280).

For detection of HCoV 229E, SuperScript III Platinum One-Step qRT-PCR kit (Invitrogen #11732088) was used. Additional MgSO_4_ was added to make a final 3.8 mM of Mg in the reaction. Reaction mixture was prepared to 23 µL and 2 µL of samples were added for a final reaction volume of 25 µL. qRT-PCR was performed using the Roche LC480 with the following condition: reverse transcription at 55 °C for 20 min, denaturation at 95 °C for 3 min, amplification of 50 cycles at 95 °C for 15 s followed by 58 °C for 30 s, and cooling at 40 °C for 30 s. qRT-PCR primers for the N-gene were 229E-N-fwd (5’-cgcaagaattcagaaccagag-3’), 229E-N-rev (5’-gggagtcaggttcttcaacaa-3’), and 229E-N-Probe (5’-6-FAM-ccacacttcaatcaaaagctcccaaatg-IBFQ-3’)^26^.

### In silico analysis of LAMP assay primers

To determine the organism exclusivity and specificity of the DW LAMP primer set, we downloaded 44,899 genomes from the NCBI Virus SARS-CoV-2 data hub (https://www.ncbi.nlm.nih.gov/labs/virus/vssi/#/) on December 9, 2020. We selected 43,849 sequences that had an identifiable N gene using the search string ATGTCTGATAATGGACCCCA from RefSeq NC_045512.2 for the SARS-CoV-2 N gene. The N gene sequences were extracted by selecting up to 1,260 bases downstream from the start of the search string using a custom python script. This subset was filtered to remove sequences containing homopolymers of three or more undetermined (“N”) bases, leaving 41,889 high-quality N gene sequences. This list was then filtered to remove sequences that were <750 bases. This requirement ensured the selected N genes would include the DW LAMP primer set. The remaining 41,858 sequences were then mapped to the the N-gene RefSeq using Geneious Prime software (Biomatters, Ltd., 2020 version) and the multiple alignment was exported as a fasta alignment file. Per-base mismatches were calculated by counting the number of exact matches at each position and subtracting from 41,858 using Linux command line utilities. Sequence homology between the DW LAMP primer set and SARS-CoV ZJ01 was determined using the “Map Primers” function in Geneious Prime, with NCBI accession AY297028 as the reference and DW LAMP primers truncated to remove the loop regions as the query. The analysis was repeated for MERS-CoV using NC_028752.1 as the reference.

### LAMP assay specificity and exclusivity

Plasmid DNA templates were acquired from Integrated DNA Technologies for MERS-CoV and SARS-CoV (#10006623 and #10006624). For SARS-CoV, MERS-CoV2 RNA standards, PCR reactions were performed using a T7 promoter containing primer sets targeting the N gene of each control plasmid as follows; SARS-T7-Fwd (5’-<u>gaaattaatacgactcactat</u>aggatgtctgataatggaccccaaaac-3’), SARS-Rev2 (5’-gaccacgtctcccaaatgcttg-3’), MERS-T7-Fwd (5’-<u>gaaattaatacgactcactata</u>ggatggcatcccctgctgcacctcg-3’), and MERS-Rev2 (5’-ggtccgcgaagaccaaaagcttg-3’). The PCR amplicons were confirmed by agarose electrophoresis and purified with DNA clean-up columns (Zymo Research). RNA was synthesized from clean PCR products using the T7 RiboMAX Express System as above. The *in vitro* transcribed RNAs were purified with Monarch RNA Cleanup Kit (NEB #T2040), aliquoted, and stored at −80 °C until used. RNA was quantified using a Nanodrop spectrophotometer, using appropriate extinction coefficients. Natrol Respiratory Panel 2 controls were purchased from Zeptometrix (#NATRPC2-BIO). RNA was extracted from these mixed organism panels using the QiAmp viral RNA Mini Kit (Qiagen #52904). Quantitative RT-PCR for the HCoV-229E N gene was performed to confirm the success of the RNA preparation from these panels, which yielded 2,030 HCoV229E copies per µl.

### Non-patient Test Samples and transport media

Normal human saliva was purchased from BioIVT from 5 males and 5 females with ages ranging from 27-42 yrs old, and a mix of races including Asian, Black, Caucasian, and Hispanic. Sputum was purchased from Discovery Life Sciences (BBL0000-AI900368151011420DD). Sputum remnant samples had been collected as potential tuberculosis diagnoses but were deemed negative. Clean Viral Transport Media (VTM) was prepared as described by the CDC (https://www.cdc.gov/coronavirus/2019-ncov/downloads/Viral-Transport-Medium.pdf). Universal Transport Media is a proprietary formulation (Copan) and was acquired and donated by Carolyn Walsh, MD. Heat inactivated SARS-CoV2 virus was purchased from ATCC or BEI (ATCC # VR-1986HK and BEI # NR-52286).

### Patient sample collection and testing

SARS-CoV-2 negative and positive nasopharyngeal swab samples were obtained by agreement from Johns Hopkins University. Samples had originally been collected in Universal Transport Media (UTM) and tested by either the NeuMoDx SARS-CoV-2 or the RealStar(R) SARS-CoV-2 RT-PCR assays ^11,27^. An aliquot of 250 μL was frozen at −80 °C after testing. Thirty SARS-CoV-2 positive samples (Ct values ranging from 14.13 to 41.35 and ten SARS-CoV-2 negative samples were included in this study.

### RPP30 and 18S rRNA sample integrity control LAMP

LAMP primers for the human RPP30 RNA were designed using the online software Primer Explorer v5 (available at http://primerexplorer.jp/e/). The RPP30 primers are RNA-specific, as they span at least one genomic intron. RPP30(13) FIP; 5’-agtttctccatggagaagcgcttttctgtttgggctctctgaa-3’, RPP30(13) BIP; 5’-tatctctacagtgaagaaacctcggttttttcttggaagctggaagac-3’, RPP30(13) F3; 5’-tgacgtggcaaatctagg-3’, RPP30(13) B3; 5’-actggggcattcttttcag-3’, RPP30(13) LF; 5’-gttggtggacaccgc-3’, RPP30(13) LB; 5’-ccatcagaaggagatgaagatt-3’. 18S rRNA primers were adapted from Lamb et al.^28^ 18S FIP; 5’-tggcctcagttccgaaaaccaattttcctggataccgcagctagg-3’, 18S BIP; 5’-ggcattcgtattgcgccgctttttggcaaatgctttcgctctg-3’, 18S F3; 5’-gttcaaagcaggcccgag-3’, 18S B3; 5’-cctccgactttcgttcttga-3’, 18S LF; 5’-agaaccgcggtcctattccattatt-3’, 18S LB; 5’-attcttggaccggcgcaag-3’. RT-LAMP assays were done using unextracted dilutions of human HEK293 cells. Cells were counted, resuspended in water and stored at −80 °C prior to direct use in LAMP reactions. RPP30 RNA amounts in HEK293 were quantified using qRT-PCR against a reference control using purified cellular RNA. For the RPP30 RNA standard, cellular RNA from HEK293T cells was extracted, cDNA was synthesized, PCR was performed with T7 promoter containing primer sets and the *in vitro* transcribed RNA was prepared as described above. Primers for the in vitro transcription amplicon are RPP30 T7 Fwd; (5’-gaaattaatacgactcactatagggtcatgggacttcagcatgg-3’) and RPP30 Rev (5’-ttaaaacaagaggaactaaagtcactt-3’). qRT-PCR primers for the RPP30 gene are RPP30q Fwd (5’-agatttggacctgcgagcg-3’), RPP30q Rev (5’-ttctgacctgaaggctctgcgcg-3’) and Rpp30 probe (5’-gagcggctgtctccacaagt [5’]FAM [3’]BHQ-1). Each HEK293 cell was determined to contain on average 9.5 copies of detectable RPP30 RNA.

## Data Availability

There are no external links to remote data. All data are included in the manuscript.

## Acknowledgments

We would like to thank Meghan Seltzer, Mike Perham, Anu Bhukel, Kristina Heiberger, Anastasia Osowski, for their logistical help enabling this work. Much of this work was funded as part of the The Tool Translation Team at the Janelia Research Campus. We benefited from discussions with the ad hoc COVID-19 Detection Team that quickly formed at Janelia that brought together many discovery scientists from many disciplines to study COVID-19 and join a global network in detecting, understanding and helping in a moment of crisis; Daryl Carson, Antony Rosen, and Hal Dietz at Johns Hopkins University for their help obtaining clinical samples used to validate our assay; Albert Yu at Brandeis, Nathan Tanner at New England Biolabs for useful discussions on LAMP methods, the GLAMP consortium, and Carolyn Walsh, MD for clinical insights and reagents.

